## Supplementary material for "Modelling Disease Mitigation at Mass Gatherings: A Case Study of COVID-19 at the 2022 FIFA World Cup": S2 Methods

### Supplementary Material: Derivation of $R_0$ Given No Population Stratification and Vaccination.

Martin David Grunnill

March 12, 2023

Model's ODEs without population stratification or vaccination:

$$
\begin{aligned}
dS/dt =& \frac{S\beta(M_H + M_I + F_I + \theta(F_A + M_A + P_A + P_I))}{N} \\
dE/dt =& \frac{S\beta(M_H + M_I + F_I + \theta(F_A + M_A + P_A + P_I))}{N} - \epsilon_1 E \\
dG_A/dt =& \epsilon_1(1 - p_s)E - \epsilon_2 G_A \\
dG_I/dt =& \epsilon_1 p_s E - \epsilon_2 G_I \\
dP_A/dt =& \epsilon_2 G_A - \epsilon_3 P_A \\
dP_I/dt =& \epsilon_2 G_I - \epsilon_3 P_I \\
dM_A/dt =& \epsilon_3 P_A - \gamma_1 M_A \\
dM_I/dt =& (1 - p_{h|s})\epsilon_3 P_I - \gamma_1 M_I \\
dM_H/dt =& p_{h|s}\epsilon_3 P_I - \epsilon_H M_H \\
dF_A/dt =& \gamma_1 M_A - \gamma_2 F_A \\
dF_I/dt =& \gamma_1 M_I - \gamma_2 F_I \\
dF_H/dt =& \epsilon_H M_H - \gamma_H F_H \\
dR/dt =& \gamma_2(F_A + F_I) + \gamma_H F_H
\end{aligned}
\tag{1}
$$

Matrix corresponding to transmission events:

$$
T = \begin{bmatrix}
0 & 0 & 0 & \beta\theta & \beta\theta & \beta\theta & \beta & \beta & \beta\theta & \beta & 0 \\
0 & 0 & 0 & 0 & 0 & 0 & 0 & 0 & 0 & 0 & 0 \\
0 & 0 & 0 & 0 & 0 & 0 & 0 & 0 & 0 & 0 & 0 \\
0 & 0 & 0 & 0 & 0 & 0 & 0 & 0 & 0 & 0 & 0 \\
0 & 0 & 0 & 0 & 0 & 0 & 0 & 0 & 0 & 0 & 0 \\
0 & 0 & 0 & 0 & 0 & 0 & 0 & 0 & 0 & 0 & 0 \\
0 & 0 & 0 & 0 & 0 & 0 & 0 & 0 & 0 & 0 & 0 \\
0 & 0 & 0 & 0 & 0 & 0 & 0 & 0 & 0 & 0 & 0 \\
0 & 0 & 0 & 0 & 0 & 0 & 0 & 0 & 0 & 0 & 0 \\
0 & 0 & 0 & 0 & 0 & 0 & 0 & 0 & 0 & 0 & 0 \\
0 & 0 & 0 & 0 & 0 & 0 & 0 & 0 & 0 & 0 & 0
\end{bmatrix}
\tag{2}
$$

Matrix corresponding to transitions between and out of infection stages:

$$
\Sigma = \begin{bmatrix}
-\epsilon_1 p_s - \epsilon_1(1 - p_s) & 0 & 0 & 0 & 0 & 0 & 0 & 0 & 0 & 0 & 0 \\
\epsilon_1(1 - p_s) & -\epsilon_2 & 0 & 0 & 0 & 0 & 0 & 0 & 0 & 0 & 0 \\
\epsilon_1 p_s & 0 & -\epsilon_2 & 0 & 0 & 0 & 0 & 0 & 0 & 0 & 0 \\
0 & \epsilon_2 & 0 & -\epsilon_3 & 0 & 0 & 0 & 0 & 0 & 0 & 0 \\
0 & 0 & \epsilon_2 & 0 & -\epsilon_3 p_{hs} - \epsilon_3(1 - p_{hs}) & 0 & 0 & 0 & 0 & 0 & 0 \\
0 & 0 & 0 & \epsilon_3 & 0 & -\gamma_{A1} & 0 & 0 & 0 & 0 & 0 \\
0 & 0 & 0 & 0 & \epsilon_3(1 - p_{hs}) & 0 & -\gamma_{I1} & 0 & 0 & 0 & 0 \\
0 & 0 & 0 & 0 & \epsilon_3 p_{hs} & 0 & 0 & -\epsilon_H & 0 & 0 & 0 \\
0 & 0 & 0 & 0 & 0 & \gamma_{A1} & 0 & 0 & -\gamma_{A2} & 0 & 0 \\
0 & 0 & 0 & 0 & 0 & 0 & \gamma_{I1} & 0 & 0 & -\gamma_{I2} & 0 \\
0 & 0 & 0 & 0 & 0 & 0 & 0 & \epsilon_H & 0 & 0 & -\gamma_H
\end{bmatrix}
\tag{3}
$$

$R_0$ is the spectral radius of Next Generation Matrix with large domain:

$$R_0 = \rho(K_L) = \rho(T(-\Sigma^{-1})) = -\frac{\beta p_{hs} p_s}{\gamma_{I2}} + \frac{\beta p_s}{\gamma_{I2}} - \frac{\beta p_{hs} p_s}{\gamma_{I1}} + \frac{\beta p_s}{\gamma_{I1}} - \frac{\beta p_s \theta}{\gamma_{A2}} + \frac{\beta \theta}{\gamma_{A2}} - \frac{\beta p_s \theta}{\gamma_{A1}} + \frac{\beta \theta}{\gamma_{A1}} + \frac{\beta p_{hs} p_s}{\epsilon_H} + \frac{\beta \theta}{\epsilon_3} \tag{4}$$

The transmission term's $\beta$ is therefore:

$$\beta = R_0 \epsilon_3 \epsilon_H \gamma_{A1} \gamma_{A2} \gamma_{I1} \gamma_{I2} \div \left( \begin{array}{c} -\epsilon_3 \epsilon_H \gamma_{A1} \gamma_{A2} \gamma_{I1} p_{hs} p_s + \epsilon_3 \epsilon_H \gamma_{A1} \gamma_{A2} \gamma_{I1} p_s - \epsilon_3 \epsilon_H \gamma_{A1} \gamma_{A2} \gamma_{I2} p_{hs} p_s + \epsilon_3 \epsilon_H \gamma_{A1} \gamma_{A2} \gamma_{I2} p_s - \\ \epsilon_3 \epsilon_H \gamma_{A1} \gamma_{I1} \gamma_{I2} p_s \theta + \epsilon_3 \epsilon_H \gamma_{A1} \gamma_{I1} \gamma_{I2} \theta - \epsilon_3 \epsilon_H \gamma_{A2} \gamma_{I1} \gamma_{I2} p_s \theta + \\ \epsilon_3 \epsilon_H \gamma_{A2} \gamma_{I1} \gamma_{I2} \theta + \epsilon_3 \gamma_{A1} \gamma_{A2} \gamma_{I1} \gamma_{I2} p_{hs} p_s + \epsilon_H \gamma_{A1} \gamma_{A2} \gamma_{I1} \gamma_{I2} \theta \end{array} \right) \tag{5}$$
