## Supplementary material for "Modelling Disease Mitigation at Mass Gatherings: A Case Study of COVID-19 at the 2022 FIFA World Cup": S1 Results

### Supplementary Material: In depth examinations of % relative differences and PRCCs.

Martin David Grunnill

March 12, 2023

#### 1 Effects of Testing Regimes

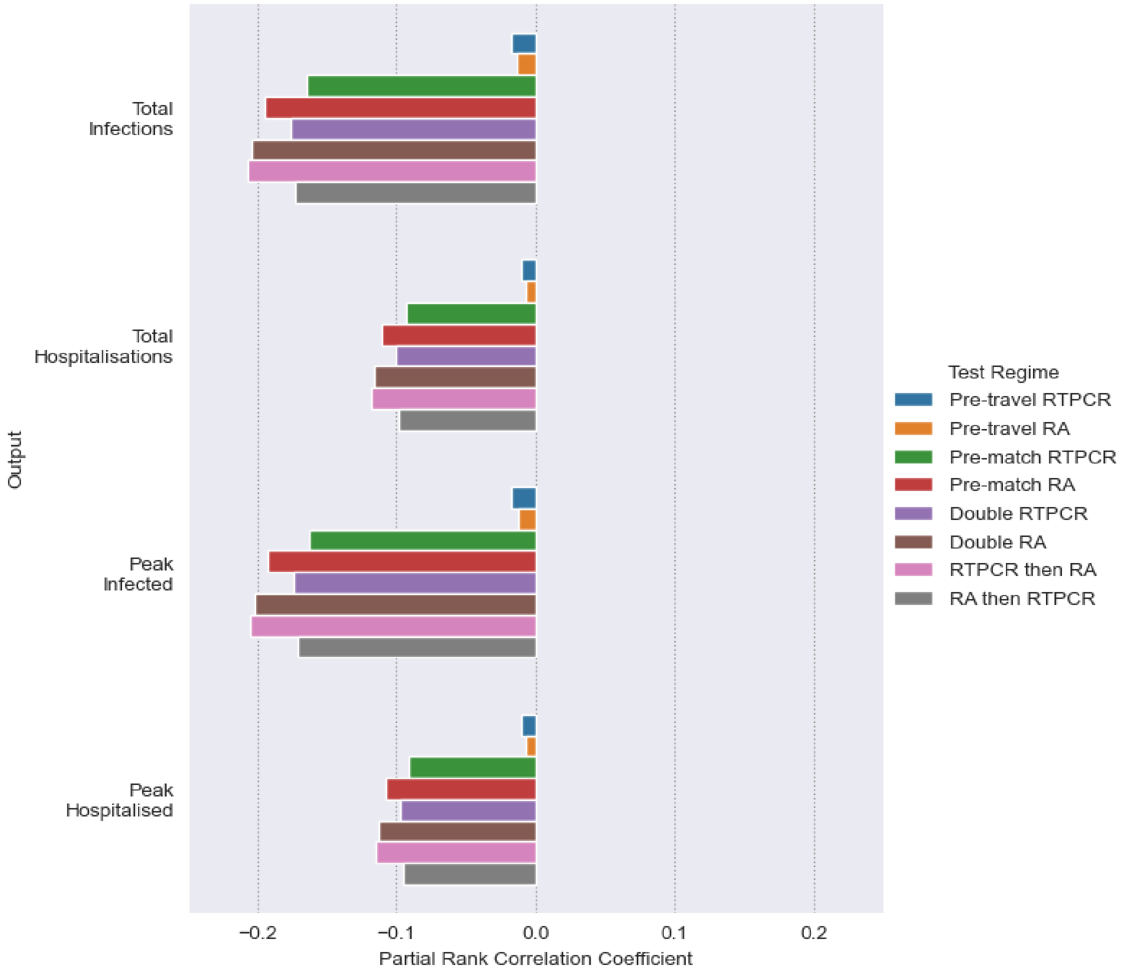


Figure 1: **Effect of different Test Regimes on infections and hospitalisations as measured by Partial Rank Correlation Coefficient (PRCC).** In calculating PRCCs Latin Hypercube (LH) sampling draws on the parameter space outlined in the main text's Tables 2, 3 and 5, using uniform distributions. Simulations are made with the resulting LH sample with each of the testing regimes outlined in the main text's Table 7. Every set of simulation made under a testing regime is given a dummy parameter value of 1, except ”No Testing” which is given a value of 0. Each testing regime's effect on an output (Total Infections, Peak Infection, Total Hospitalisation or Peak Hospitalisation) is measured through calculating PRCCs using the dummy parameter comparing the 1 for the particular testing regime and 0 for its absence.

From Figures 1 and 2 can be seen that PRCCs and % Relative Differences comparing testing regimes to peak infections and hospitalisations follow similar trends to total infections and hospitalisations.

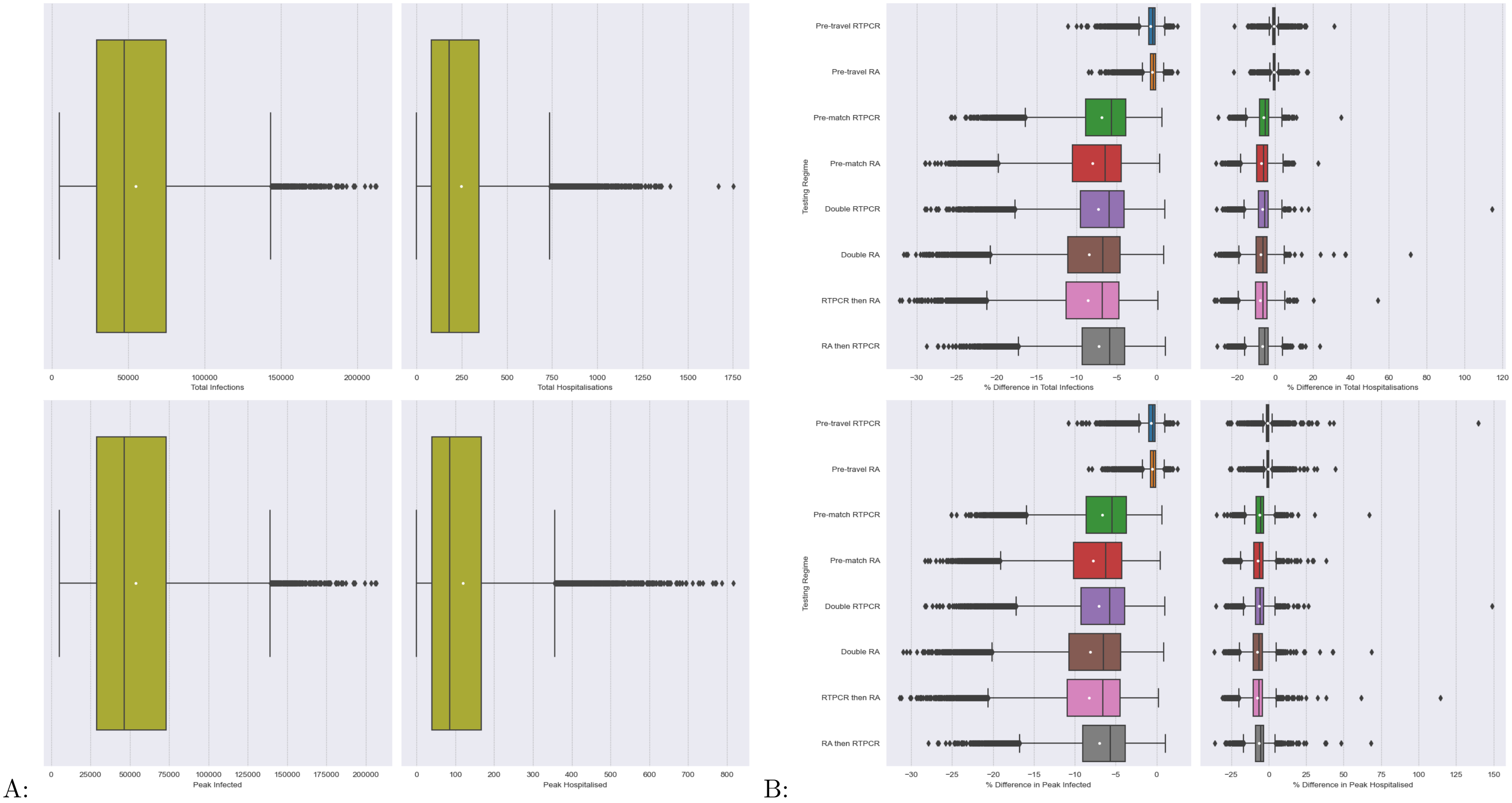


Figure 2: **Effect of different Test Regimes on infections and hospitalisations as measured by % Relative Difference to simulations with no testing regime. A: Boxplots Total Infections and Hospitalisation in simulations made with no testing regime. B: Boxplots of a Testing Regimes % Relative Differences in Total infections and Hospitalisation.** For every parameter set produced under LHS the % relative difference in outputs simulated under a testing regime, Fig B, was calculated against the corresponding output from the "No Testing" regime simulations, depicted in Fig A, as a control (see main text's Eq 4). The white dots are the means. The array of samples used in simulation was generated from Latin Hypercube sampling drawing upon the distributions outlined in the main text's Tables 2, 3 and 5. Details of testing regimes can be found in the main text's Table 7.

#### 2 Effects of Starting Conditions

Increasing the number of attendees, $N_A$ at match has very strong correlation with both infections and hospitalisations (see Figure 3). This is mitigated by increasing the proportion of match attendees that are from the host nation $N_Q^*$, thereby decreasing the number of seed infections within visitor cluster's A and B. With regards to both hospitalisations and infections it can be seen that both of their totals and peaks follow similar patterns of correlation with regards to the models starting conditions

#### 3 Effects of Parameters Related to Disease Process

Figure 4 demonstrates that increasing $R_0$ and to a lesser extent the transmission on match day $b$ leads to rapid increases in infections and hospitalisations This can be mitigated by improving the efficacy of those in the effectively vaccinated group in terms of decreasing susceptibility to infection ($l_E$). Increasing the efficacy of vaccination against hospitalisation for this group ($VE_H$) decreases hospitalisations but has little effect on infections. Rate of being hospitalised ($\epsilon_h$) and recovery from being hospitalised ($\gamma_h$) have little effect on outputs other than Peak Hospitalised. $\epsilon_h$ increasing the peak in hospitalised and $_h$ reducing the peak. Unsurprisingly increasing the proportion of those symptomatic who become hospitalised ($p_{h|s}$) leads to dramatic increases in hospitalisation.

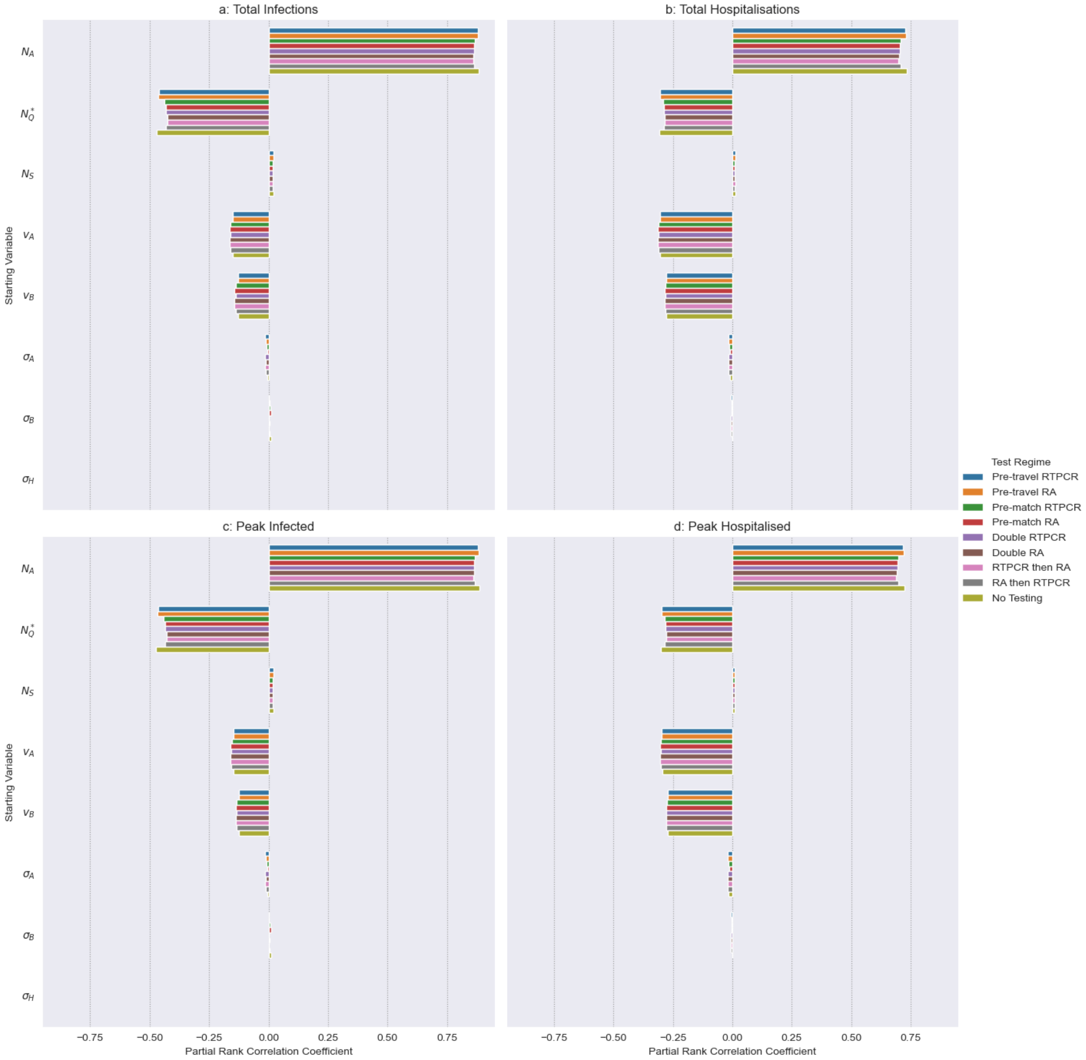


Figure 3: **Partial Rank Correlation Coefficients between starting conditions and Total infections (a), Total Hospitalisation (b), Peak infections (c) and Peak Hospitalisation (d).** $N_A$ number of match attendees (4000-80000), $N_Q^*$ proportion of tickets given to Qatari's (0-0.5). $N_S$ number of staff (4000-20000). $v_A$ proportion recently vaccinated in visitor group cluster A (0-1). $v_B$ proportion recently vaccinated in visitor group cluster B (0-1). $\sigma_A$ or $\sigma_B$ prevalence of visitor cluster A and B, respectively ($4.47 \times 10^{-6}$ to 0.003). $\sigma_H$ prevalence of host clusters (0.0006 to 0.0011). The array of samples used in simulation was generated from Latin Hypercube sampling drawing upon the distributions outlined above and in the main text's Tables 2, 3 and 5, using uniform distributions. Details of testing regimes can be found in the main text's Table 7.

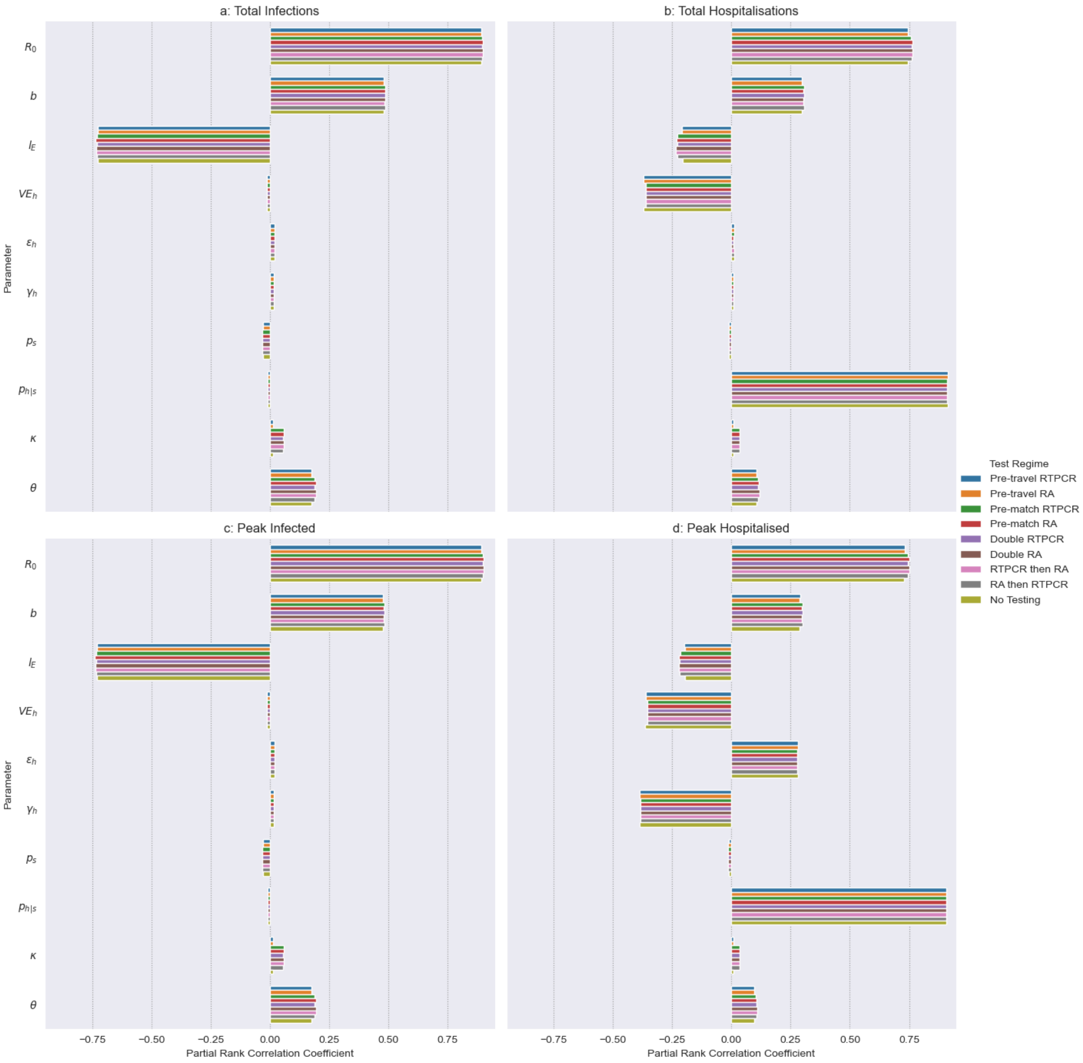


Figure 4: **Partial Rank Correlation Coefficients comparing parameters effects on Total infections (a), Total Hospitalisation (b), Peak infections (c) and Peak Hospitalisation (d).** $R_0$ basic reproduction number (2-7). $b$ increase in Transmission Match Day (1-78.5). $l_E$ recent vaccine efficacy against infection (0.173-0.775). $VE_h$ recent vaccine efficacy against hospitalising infection (0.837-1). $\epsilon_h$ rate of hospitalisation (0.103-0.382). $\gamma_h$ rate of hospital recovery (0.0448-0.155). $p_s$ proportion symptomatic (0.41-0.84). $p_{h|s}$ proportion hospitalised given being symptomatic (0-0.0234). $\kappa$ isolation transmission modifier (0-1). $\theta$ asymptomatic transmission modifier (0.342-1). The array of samples used in simulation was generated from Latin Hypercube sampling drawing upon the distributions outlined above and in the main text's Tables 2, 3 and 5, using uniform distributions. Details of testing regimes can be found in the main text's Table 7.
