## Supplementary material for "Modelling Disease Mitigation at Mass Gatherings: A Case Study of COVID-19 at the 2022 FIFA World Cup": S1 Table

### Supplementary Material: Differences in PRCCs between proportions of visitor clusters A and B effectively vaccinated ($v_A$ and $v_B$) and different testing regimes.

Martin David Grunnill

March 12, 2023

Table 1: **Differences in PRCCs between different testing regimes and proportions of visitor clusters A effectively vaccinated ($v_A$) under the "No Testing" test regime.**

| Test Regime | Output | Differences in PCC | z-score | p_value (one tailed) | p_value (two tailed) |
|---|---|---|---|---|---|
| Pre-travel RTPCR | peak hospitalised | 0.285485156 | 23.29013569 | 2.79E-120 | 5.58E-120 |
| Pre-travel RTPCR | peak infected | 0.129090405 | 10.53131131 | 3.10E-26 | 6.20E-26 |
| Pre-travel RTPCR | total hospitalisations | 0.29415326 | 23.99728747 | 1.48E-127 | 2.97E-127 |
| Pre-travel RTPCR | total infections | 0.133077878 | 10.85661291 | 9.28E-28 | 1.86E-27 |
| Pre-travel RA | peak hospitalised | 0.288110701 | 23.50433005 | 1.84E-122 | 3.68E-122 |
| Pre-travel RA | peak infected | 0.133793811 | 10.91501936 | 4.89E-28 | 9.77E-28 |
| Pre-travel RA | total hospitalisations | 0.296710205 | 24.20588532 | 9.64E-130 | 1.93E-129 |
| Pre-travel RA | total infections | 0.137796267 | 11.2415434 | 1.27E-29 | 2.55E-29 |
| Pre-match RTPCR | peak hospitalised | 0.204216058 | 16.66012959 | 1.28E-62 | 2.55E-62 |
| Pre-match RTPCR | peak infected | -0.016631435 | -1.356807408 | 0.087421207 | 0.174842413 |
| Pre-match RTPCR | total hospitalisations | 0.210622949 | 17.18280958 | 1.79E-66 | 3.57E-66 |
| Pre-match RTPCR | total infections | -0.014160842 | -1.155254258 | 0.123993161 | 0.247986321 |
| Pre-match RA | peak hospitalised | 0.187966736 | 15.3344953 | 2.25E-53 | 4.50E-53 |
| Pre-match RA | peak infected | -0.046151116 | -3.765049563 | 8.33E-05 | 0.000166516 |
| Pre-match RA | total hospitalisations | 0.1937887 | 15.80945636 | 1.34E-56 | 2.68E-56 |
| Pre-match RA | total infections | -0.044061205 | -3.594552704 | 0.000162475 | 0.000324949 |
| Double RTPCR | peak hospitalised | 0.197806124 | 16.13720145 | 6.99E-59 | 1.40E-58 |
| Double RTPCR | peak infected | -0.02778936 | -2.267080963 | 0.01169264 | 0.02338528 |
| Double RTPCR | total hospitalisations | 0.203861313 | 16.6311892 | 2.07E-62 | 4.14E-62 |
| Double RTPCR | total infections | -0.025474225 | -2.078210188 | 0.018845002 | 0.037690004 |
| Double RA | peak hospitalised | 0.182140341 | 14.85917278 | 3.03E-50 | 6.07E-50 |
| Double RA | peak infected | -0.055514067 | -4.528887535 | 2.96E-06 | 5.93E-06 |
| Double RA | total hospitalisations | 0.187965284 | 15.33437687 | 2.25E-53 | 4.51E-53 |
| Double RA | total infections | -0.053584406 | -4.371463987 | 6.17E-06 | 1.23E-05 |
| RTPCR then RA | peak hospitalised | 0.180141577 | 14.69611179 | 3.41E-49 | 6.83E-49 |
| RTPCR then RA | peak infected | -0.058808634 | -4.797661221 | 8.03E-07 | 1.61E-06 |
| RTPCR then RA | total hospitalisations | 0.185887191 | 15.16484415 | 3.02E-52 | 6.04E-52 |
| RTPCR then RA | total infections | -0.056910348 | -4.642797346 | 1.72E-06 | 3.44E-06 |
| RA then RTPCR | peak hospitalised | 0.199562542 | 16.2804916 | 6.79E-60 | 1.36E-59 |
| RA then RTPCR | peak infected | -0.024925421 | -2.033438217 | 0.021004135 | 0.04200827 |
| RA then RTPCR | total hospitalisations | 0.205783196 | 16.7879782 | 1.49E-63 | 2.99E-63 |
| RA then RTPCR | total infections | -0.02258497 | -1.8425022 | 0.032700863 | 0.065401725 |

Table 2: **Differences in PRCCs between different testing regimes and proportions of visitor clusters B effectively vaccinated ($v_B$) under the "No Testing" test regime.**

| Test Regime | Output | Differences in PCC | z-score | p_value (one tailed) | p_value (two tailed) |
|---|---|---|---|---|---|
| Pre-travel RTPCR | peak hospitalised | 0.260997181 | 21.29238473 | 6.68E-101 | 1.34E-100 |
| Pre-travel RTPCR | peak infected | 0.10663991 | 8.699779777 | 1.66E-18 | 3.33E-18 |
| Pre-travel RTPCR | total hospitalisations | 0.266416197 | 21.73447292 | 4.84E-105 | 9.69E-105 |
| Pre-travel RTPCR | total infections | 0.11089937 | 9.047270284 | 7.33E-20 | 1.47E-19 |
| Pre-travel RA | peak hospitalised | 0.263622727 | 21.50657909 | 6.76E-103 | 1.35E-102 |
| Pre-travel RA | peak infected | 0.111343316 | 9.083487829 | 5.26E-20 | 1.05E-19 |
| Pre-travel RA | total hospitalisations | 0.268973142 | 21.94307077 | 5.04E-107 | 1.01E-106 |
| Pre-travel RA | total infections | 0.115617759 | 9.432200771 | 2.01E-21 | 4.02E-21 |
| Pre-match RTPCR | peak hospitalised | 0.179728083 | 14.66237863 | 5.61E-49 | 1.12E-48 |
| Pre-match RTPCR | peak infected | -0.039081929 | -3.18833894 | 0.000715463 | 0.001430927 |
| Pre-match RTPCR | total hospitalisations | 0.182885886 | 14.91999504 | 1.22E-50 | 2.44E-50 |
| Pre-match RTPCR | total infections | -0.03633935 | -2.964596884 | 0.001515399 | 0.003030798 |
| Pre-match RA | peak hospitalised | 0.163478761 | 13.33674434 | 7.08E-41 | 1.42E-40 |
| Pre-match RA | peak infected | -0.068601611 | -5.596581096 | 1.09E-08 | 2.19E-08 |
| Pre-match RA | total hospitalisations | 0.166051637 | 13.54664181 | 4.15E-42 | 8.29E-42 |
| Pre-match RA | total infections | -0.066239713 | -5.40389533 | 3.26E-08 | 6.52E-08 |
| Double RTPCR | peak hospitalised | 0.17331815 | 14.13945049 | 1.08E-45 | 2.17E-45 |
| Double RTPCR | peak infected | -0.050239854 | -4.098612496 | 2.08E-05 | 4.16E-05 |
| Double RTPCR | total hospitalisations | 0.17612425 | 14.36837465 | 4.09E-47 | 8.17E-47 |
| Double RTPCR | total infections | -0.047652733 | -3.887552814 | 5.06E-05 | 0.00010126 |
| Double RA | peak hospitalised | 0.157652367 | 12.86142182 | 3.71E-38 | 7.42E-38 |
| Double RA | peak infected | -0.077964562 | -6.360419068 | 1.01E-10 | 2.01E-10 |
| Double RA | total hospitalisations | 0.160228221 | 13.07156232 | 2.39E-39 | 4.79E-39 |
| Double RA | total infections | -0.075762914 | -6.180806613 | 3.19E-10 | 6.38E-10 |
| RTPCR then RA | peak hospitalised | 0.155653602 | 12.69836083 | 3.02E-37 | 6.04E-37 |
| RTPCR then RA | peak infected | -0.081259128 | -6.629192753 | 1.69E-11 | 3.38E-11 |
| RTPCR then RA | total hospitalisations | 0.158150128 | 12.9020296 | 2.19E-38 | 4.38E-38 |
| RTPCR then RA | total infections | -0.079088856 | -6.452139972 | 5.51E-11 | 1.10E-10 |
| RA then RTPCR | peak hospitalised | 0.175074568 | 14.28274064 | 1.40E-46 | 2.80E-46 |
| RA then RTPCR | peak infected | -0.047375915 | -3.86496975 | 5.56E-05 | 0.000111103 |
| RA then RTPCR | total hospitalisations | 0.178046133 | 14.52516365 | 4.20E-48 | 8.39E-48 |
| RA then RTPCR | total infections | -0.044763479 | -3.651844825 | 0.000130182 | 0.000260363 |
